# Post-vaccination SARS-CoV-2 infection: risk factors and illness profile in a prospective, observational community-based case-control study

**DOI:** 10.1101/2021.05.24.21257738

**Authors:** Michela Antonelli, Rose S Penfold, Jordi Merino, Carole H Sudre, Erika Molteni, Sarah Berry, Liane S Canas, Mark S Graham, Kerstin Klaser, Marc Modat, Benjamin Murray, Eric Kerfoot, Liyuan Chen, Jie Deng, Marc F Österdahl, Nathan J Cheetham, David Drew, Long Nguyen, Joan Capdevila Pujol, Christina Hu, Somesh Selvachandran, Lorenzo Polidori, Anna May, Jonathan Wolf, Andrew T Chan, Alexander Hammers, Emma L Duncan, Tim D Spector, Sebastien Ourselin, Claire J Steves

## Abstract

**Background:** COVID-19 vaccines show excellent efficacy in clinical trials and real-world data, but some people still contract SARS-CoV-2 despite vaccination. This study sought to identify risk factors associated with SARS-CoV-2 infection post-vaccination and describe characteristics of post-vaccination illness.

**Methods:** Amongst 1,102,192 vaccinated UK adults from the COVID Symptom Study, 2394 (0.2%) cases of post-vaccination SARS-CoV-2 infection were identified between 8th December 2020 and 1st May 2021. Using a control group of vaccinated individuals testing negative, we assessed the associations of age, frailty, comorbidity, area-level deprivation and lifestyle factors with infection. Illness profile post-vaccination was assessed using a second control group of unvaccinated cases.

**Findings:** Older adults with frailty (OR=2.78, 95% CI=[1.98-3.89], p-value<0.0001) and individuals living in most deprived areas (OR=1.22 vs. intermediate group, CI[1.04-1.43], p-value=0.01) had increased odds of post-vaccination infection. Risk was lower in individuals without obesity (OR=0.6, CI[0.44-0.82], p-value=0.001) and those reporting healthier diet (OR=0.73, CI[0.62-0.86], p-value<0.0001). Vaccination was associated with reduced odds of hospitalisation (OR=0.36, CI[0.28-0.46], p-value<0.0001), and high acute-symptom burden (OR=0.51, CI[0.42-0.61], p-value<0.0001). In older adults, risk of ≥28 days illness was lower following vaccination (OR=0.72, CI[0.51-1.00], p-value=0.05). Symptoms were reported less in positive-vaccinated vs. positive-unvaccinated individuals, except sneezing, which was more common post-vaccination (OR=1.24, CI[1.05-1.46], p-value=0.01).

**Interpretation:** Our findings suggest that older individuals with frailty and those living in most deprived areas are at increased risk of infection post-vaccination. We also showed reduced symptom burden and duration in those infected post-vaccination. Efforts to boost vaccine effectiveness in at-risk populations, and to targeted infection control measures, may still be appropriate to minimise SARS-CoV-2 infection.

**Funding:** This work is supported by UK Department of Health via the National Institute for Health Research (NIHR) comprehensive Biomedical Research Centre (BRC) award to Guy’s & St Thomas’ NHS Foundation Trust in partnership with King’s College London and King’s College Hospital NHS Foundation Trust and via a grant to ZOE Global; the Wellcome Engineering and Physical Sciences Research Council (EPSRC) Centre for Medical Engineering at King’s College London (WT 203148/Z/16/Z). Investigators also received support from the Chronic Disease Research Foundation, the Medical Research Council (MRC), British Heart Foundation, the UK Research and Innovation London Medical Imaging & Artificial Intelligence Centre for Value Based Healthcare, the Wellcome Flagship Programme (WT213038/Z/18/Z and Alzheimer’s Society (AS-JF-17-011), and the Massachusetts Consortium on Pathogen Readiness (MassCPR).

**Research in context:** *Evidence before this study:* To identify existing evidence for risk factors and characteristics of SARS-CoV-2 infection post-vaccination, we searched PubMed for peer-reviewed articles published between December 1, 2020 and May 18, 2021 using keywords (“COVID-19” OR “SARS-CoV-2”) AND (“Vaccine” OR “vaccination”) AND (“infection”) AND (“risk factor*” OR “characteristic*”). We did not restrict our search by language or type of publication. Of 202 articles identified, we found no original studies on individual risk and protective factors for COVID-19 infection following vaccination nor on nature and duration of symptoms in vaccinated, community-based individuals. Previous studies in unvaccinated populations have shown that social and occupational factors influence risk of SARS-CoV-2infection, and that personal factors (age, male sex, multiple morbidities and frailty) increased risk for adverse outcomes in COVID-19. Phase III clinical trials have demonstrated good efficacy of BNT162b2 and ChAdOx1 vaccines against SARS-CoV-2 infection, confirmed in published real-world data, which additionally showed reduced risk of adverse outcomes including hospitalisation and death.

*Added value of this study:* This is the first observational study investigating characteristics of and factors associated with SARS-CoV-2 infection after COVID-19 vaccination. We found that vaccinated individuals with frailty had higher rates of infection after vaccination than those without. Adverse determinants of health such as increased social deprivation, obesity, or a less healthy diet were associated with higher likelihood of infection after vaccination. In comparison with unvaccinated individuals, those with post-vaccination infection had fewer symptoms of COVID-19, and more were entirely asymptomatic. Fewer vaccinated individuals experienced five or more symptoms, required hospitalisation, and, in the older adult group, fewer had prolonged illness duration (symptoms lasting longer than 28 days).

*Implications of all the available evidence:* Some individuals still contract COVID-19 after vaccination and our data suggest that frail older adults and those living in more deprived areas are at higher risk. However, in most individuals illness appears less severe, with reduced need for hospitalisation and lower risk of prolonged illness duration. Our results are relevant for health policy post-vaccination and highlight the need to prioritise those most at risk, whilst also emphasising the balance between the importance of personal protective measures versus adverse effects from ongoing social restrictions. Strategies such as timely prioritisation of booster vaccination and optimised infection control could be considered for at-risk groups. Research is also needed on how to enhance the immune response to vaccination in those at higher risk.

## Introduction

Vaccination against SARS-CoV-2 is a leading strategy to change the course of the pandemic worldwide. The United Kingdom was the first country internationally to authorise a vaccine against SARS-CoV-2,^1^ with three currently licensed: BNT162b2 (“Pfizer-BioNTech”), mRNA-1273 (“Moderna”) and ChAdOx1 nCoV-19 (“Oxford-AstraZeneca”), each with good efficacy in Phase 3 clinical trials (2-5). As of 14 May 2021, ∼36.3 million (69%) of the UK adult population had received at least one vaccination (6). UK data presents an early window on real-world efficacy and also on the remaining challenges post-vaccination.

Previous analysis of community-based individuals using the COVID Symptom Study showed significant reduction in infection post-vaccination from 12 days after first vaccine dose (7), findings recapitulated in a UK-based “real world” case-control study (8). National surveillance data from the first four months of Israel’s vaccination campaign showed two doses of BNT162b2 prevented symptomatic and asymptomatic infections, COVID-19-related hospitalisations, severe disease, and death (9).

None-the-less, some still contract COVID-19 after vaccination, and further virus variants may evolve with increased transmissibility (as with B.1.1.7 and B.1.617.2) (10, 11). Early data suggest that while COVID-19 is usually milder if contracted post-vaccination, mortality remains high in hospitalised individuals: Recent International Severe Acute Respiratory and Emerging Infection Consortium (ISARIC) data showed mortality of 28% in individuals hospitalised with COVID-19 >21 days post-vaccination, similar to mortality rates during the first wave (March-April 2020) (12, 13).

Identifying and protecting individuals at higher risk of post-vaccination infection will become increasingly salient as more of the population is vaccinated. Groups at higher risk of SARS-CoV-2 infection before vaccine availability included frontline healthcare workers and individuals from areas of greater relative deprivation (likely reflecting increased exposure) (14, 15), and increasing age, male sex, multi-morbidity and frailty are associated with poorer COVID-19 outcomes (16-18). However, to our knowledge there are no studies investigating risk factors for post-vaccination infection.

Individuals with COVID-19 have differing symptoms and clinical needs (19). Elucidating symptom profiles in individuals with COVID-19 post-vaccination has clinical utility, facilitating identification of risk groups for intervention, predicting medical resource requirements and informing appropriate testing guidelines. Finally, some unvaccinated individuals with COVID-19 experience prolonged illness duration (‘Long-COVID’) (20). Whether this risk is similar in individuals infected post-vaccination is unknown.

This study aimed to:

1. Describe individual factors associated with SARS-CoV-2 infection at least 14 days after first vaccination,
2. Assess illness duration, severity, and symptom profile in individuals with SARS-CoV-2 infection after first vaccination compared to unvaccinated individuals with SARS-CoV-2 infection.

## Methods

### Study design and participants

This community-based case-control study used data from the COVID Symptom Study logged through a free smartphone app developed by Zoe Global (London, UK) and King’s College London (London, UK) (21). The app was launched in the UK on 24 March 2020, with nearly 4.5 million unique participants providing data by self-or proxy-report. At registration, each participant reported baseline demographic information (e.g., age, sex, ethnicity, whether a healthcare worker) geographic location, and information on health risk factors including comorbitidies, lifestyle, frailty and visits to hospital. Participants were encouraged to self-report any pre-specified symptoms daily, enabling prospective,longitudinal information on incident symptoms. Those experiencing new symptoms were invited for a SARS-CoV-2 test through local testing centres. All users were prompted to record any COVID-19 testing results (whether prompted by the app or for other reasons), and from December 2020, any COVID-19 vaccine(s) and subsequent symptoms (21).

In this nested case-control study, inclusion criteria for cases were: 1) age ≥18 years; 2) living in the UK; 3) first dose of a COVID-19 vaccine between 8 December 2020 and 1 May 2021; 4) at least 14 days of app usage after vaccination; 5) a positive RT-PCR or lateral flow antigen test (LFAT) at least fourteen days after first vaccination, but before the second dose (if more than one test result reported, only the first positive test was selected); 6) no positive SARS-CoV-2 test prior to vaccination.

To identify risk factors for post-vaccination infection, we selected controls among vaccinated UK adults reporting negative RT-PCR or LFAT before second vaccination, and until 14 May 2021 (date of data extraction) (CG-1), matching 1:1 with cases for date of post-vaccination test, healthcare worker status, and sex. If multiple negative tests were reported, the last test date was used for matching. To compare symptoms of SARS-CoV-2 infection pre- and post-vaccination, we un-vaccinated selected controls aged ≥18 years, living in the UK, who reported a positive SARS-CoV-2 test, regardless of symptoms (CG-2). Controls were matched 1:1 with cases using the date of positive test, healthcare worker status, sex, BMI, and age. For both control groups we used a matching algorithm based on minimum Euclidean distance (22) between the vectors of the covariates, with sex as a binary variable multiplied by 100 to ensure balance between covariate strengths.

Data from 1,531,762 app users reporting a RT-PCR or LFAT test within the study period were processed to obtain weights for inverse probability of being vaccinated (IPW).

### Risk factor variable definitions

For this analysis the outcome variable was case status (self-reported positive RT-PCR test or LFAT for SARS-CoV-2). We considered a-priori defined risk factors for SARS-CoV-2 infection based on previous evidence for unvaccinated individuals(15-17): age; BMI; self-reported comorbidities including cancer, diabetes, asthma, lung disease, heart disease, and kidney disease analysed individually as binary variables; dependency level (frailty), assessed by the PRISMA7 questionnaire (embedded in app registration) (23, 24), as a binary variable (PRISMA7 ≥ 3 = frail; PRISMA7 < 3 = not frail) (25); local area Index of Multiple Deprivation (IMD), a score, ranging from 1 (most deprived) to 10 (least deprived) estimating relative locality deprivation derived from postal code, divided into low IMD[1-3], middle IMD[4-7], and high IMD[8-10] groups (26); four healthy lifestyle factors including no current smoking, no obesity (Body Mass Index<30), physical activity at least once weekly, and a healthier diet pattern. We also considered a healthy lifestyle score based on these four healthy lifestyle factors (27) (see Supplementary Methods).

### Disease severity and symptom definitions

To compare COVID-19 symptoms and severity outcomes in vaccinated vs. unvaccinated individuals testing positive for SARS-CoV-2, we assessed: 1) disease severity, assessed as: asymptomatic/symptomatic; >5 symptoms/≤5 symptoms reported in the first week of illness (19); and self-reported presentation to hospital/no hospital presentation; 2) illness duration, assessed as duration <28days/ duration≥28days; 3) individual symptom reports. Vaccination status was the exposure. For cases and CG-2, symptoms were considered within a window from three days before and up to fourteen days after the test date for SARS-CoV-2 (see Supplementary Table 1 for complete list).

### Statistical analyses

Data census was 14 May 2021. Data were extracted and preprocessed using ExeTera13, a Python library developed at KCL (28), and openly available on GitHub. Statistical analysis was run using Python 3.7 and the following packages: numpy v1.19.2, pandas v1.1.3, scipy 1.5.2, and statsmodels v0.12.1.

#### Risk factor analysis

Differences in proportions and means of covariates between cases and respective controls were assessed using Fisher’s exact test for categorical variables and Wilcoxon’s test for continuous variables. Univariate logistic regression models (adjusted for age, BMI, and sex) were used to analyse association between risk factor variables and post-vaccination infection. As factors associated may differ by age group, all analyses were stratified by sex and age (younger adults: 18-59 years; older adults, ≥60 years). Multivariable logistic regression was then used to assess independence of the variables.

To examine whether health-conscious behaviours might explain the association of lifestyle factors and infection post-vaccination, we adjusted models for reported individual adherence to mask-wearing guidance during 2020. Finally, we examined models using inverse probability weighting (IPW) (29) to check for potential index event bias of vaccination using weights derived from probabilities of being vaccinated in the population tested and active on the app during the study period (Supplementary Figure 1).

#### Analyses of reported symptoms

Univariate logistic regression models adjusted by age, BMI, and sex were used to assess association of individual symptoms, overall illness duration, and disease severity (outcomes), with vaccination status (exposure). Symptoms were examined if reported by >1% of app users reporting a positive test. We provide models additionally adjusting for frailty and comorbidities, given their association with the exposure (vaccination) and outcome (symptoms) which may confound any observed associations.

This study reports on BNT162b2 and ChAdOx1 vaccines only, as there were no positive cases who received the mRNA-1273 vaccine. Post-vaccination infection was similar in these two vaccine types therefore combined analysis was performed.

### Ethical approval

All app users provided informed consent for data usage for COVID-19-related research. In the UK, the app and study were approved by King’s College London (KCL) ethics committee (REMAS no. 18210, reference LRS-19/20–18210).

## Results

Between 8 December 2020 and 14 May 2021, 1,102,192 app users reported a first dose and 559,962 a second dose of a COVID-19 vaccine (approximately one-third BNT162b2, two-thirds ChAdOx1). Of these, 2,394 (0.2%) and 187 (0.03%) reported testing positive for SARS-CoV-2 at least 14 days after first and second vaccination respectively. Supplementary Figure 2 shows time in days from the first or second dose and day of positive test which also reflects changing incidence of COVID-19 infection in the UK population (30).

### Risk/protective factor analysis

Table 1 shows demographic information of cases and controls for risk factor analysis stratified by age group. 69.5% of participants were female. Cases were significantly younger (p-value< 0.0001) and had higher BMI (p-value< 0.0001). Asthma and lung disease were the most commonly reported comorbidities in both cases and controls; there was no significant difference in prevalence of comorbidities between cases and controls. Cases were more likely to smoke, have a sedentary lifestyle, and less healthy diet.

**Table 1.** Demographics of post-vaccination cases and controls for risk factor analysis.

|  | Positive group |  |  | Control group 1: vaccinated users testing negative, matched on healthcare worker status, sex and day of test |  |  |
| --- | --- | --- | --- | --- | --- | --- |
|  | All<br>n=2394 | Younger adults<br>(18-59 years)<br>n=1372 | Older adults<br>(60+ years)<br>n=1022 | All<br>n=2394 | Younger adults<br>(18-59 years)<br>n=1467 | Older adults<br>(60+ years)<br>n=927 |
| **Basic characteristics** |  |  |  |  |  |  |
| Female n(%) | 1663(69.5) | 1037(75.6) | 626(61.3) | 1663(69.5)<sup>=</sup> | 1136(77.4)<sup>=</sup> | 527(56.9)<sup>*</sup> |
| Age (SD) | 52.9(15.1) | 42.7(9.1) | 66.6(9.8) | 54.8(15.5)<sup>*</sup> | 45.0(10.2)<sup>*</sup> | 70.3(7.9)<sup>*</sup> |
| BMI (SD) | 27.6(7.0) | 27.9(7.3) | 27.2(6.6) | 26.7(6.8)<sup>*</sup> | 26.9(7.1)<sup>*</sup> | 26.3(6.4)<sup>*</sup> |
| HCW n(%) | 643(26.9) | 459(33.5) | 184(18.0) | 642(26.8)<sup>=</sup> | 561(38.2)<sup>*</sup> | 81(8.7)<sup>*</sup> |
| **Vaccine type** |  |  |  |  |  |  |
| BNT162b2 | 1427(59.6) | 871(63.5) | 556(54.4) | 1477(61.7)<sup>=</sup> | 951(64.8)<sup>=</sup> | 526(56.7)<sup>=</sup> |
| ChAdOx1 | 888(37.1) | 446(32.5) | 442(43.2) | 762(31.8)<sup>*</sup> | 384(26.2)<sup>*</sup> | 378(40.8)<sup>=</sup> |
| Not sure | 79(3.3) | 55(4.0) | 24(2.3) | 155(6.5)<sup>*</sup> | 132(9.0)<sup>*</sup> | 23(2.5)<sup>=</sup> |
| **Comorbidities** |  |  |  |  |  |  |
| Cancer n(%) | 45(1.9) | 7(0.5) | 38(3.7) | 51(2.1)<sup>=</sup> | 10(0.7)<sup>=</sup> | 41(4.4)<sup>=</sup> |
| Diabetes n(%) | 97(4.1) | 26(1.9) | 71(6.9) | 93(3.9)<sup>=</sup> | 36(2.5)<sup>=</sup> | 57(6.1)<sup>=</sup> |
| Lung disease n(%) | 261(10.9) | 148(10.8) | 113(11.1) | 258(10.8)<sup>=</sup> | 180(12.3)<sup>=</sup> | 78(8.4)<sup>*</sup> |
| Heart disease n(%) | 119(5.0) | 17(1.2) | 102(10.0) | 124(5.2)<sup>=</sup> | 24(1.6)<sup>=</sup> | 100(10.8)<sup>=</sup> |
| Kidney disease n(%) | 35(1.5) | 13(0.9) | 22(2.2) | 29(1.2)<sup>=</sup> | 14(1.0)<sup>=</sup> | 15(1.6)<sup>=</sup> |
| Asthma n(%) | 348(14.5) | 216(15.7) | 132(12.9) | 342(14.3)<sup>=</sup> | 246(16.8)<sup>=</sup> | 96(10.4)<sup>=</sup> |
| Frailty n (%) | 162(6.8) | 24(1.7) | 138(13.5) | 159(6.6)<sup>=</sup> | 41(2.8)<sup>=</sup> | 118(12.7)<sup>=</sup> |
| Comorbidity status | 592(24.7) | 280(20.4) | 312(30.5) | 604(25.2)<sup>=</sup> | 322(21.9)<sup>=</sup> | 282(30.4)<sup>=</sup> |
| **IMD** |  |  |  |  |  |  |
| IMD [1-3] n(%) | 528(22.1) | 316(23.0) | 212(20.7) | 415(17.3)<sup>*</sup> | 267(18.2)<sup>*</sup> | 148(16.0)<sup>*</sup> |
| IMD [4-7] n(%) | 947(39.6) | 566(41.3) | 381(37.3) | 921(38.5)<sup>=</sup> | 582(39.7)<sup>=</sup> | 339(36.6)<sup>=</sup> |
| IMD [8-10] n(%) | 919(38.4) | 490(35.7) | 429(42.0) | 1058(44.2)<sup>*</sup> | 618(42.1)<sup>*</sup> | 440(47.5)<sup>*</sup> |
| **Healthy lifestyle<sup>1</sup>** |  |  |  |  |  |  |
| No smoking n(%) | 1211(97.1) | 863(96.5) | 339(98.5) | 1226(98.3)<sup>=</sup> | 701(97.6)<sup>=</sup> | 516(99.2)<sup>=</sup> |
| Not obese n(%) | 886(71.1) | 621(69.5) | 265(77.0) | 1007(80.8)<sup>*</sup> | 564(78.6)<sup>*</sup> | 443(85.2)<sup>*</sup> |
| Healthier diet n(%) | 422(33.8) | 276(30.9) | 140(40.7) | 545(43.7)<sup>*</sup> | 282(39.3)<sup>*</sup> | 261(50.2)<sup>*</sup> |
| Not sedentary n(%) | 952(76.3) | 681(76.2) | 265(77.0) | 982(78.7)<sup>=</sup> | 570(79.4)<sup>=</sup> | 406(78.1)<sup>=</sup> |
| Healthy lifestyle score | 2.8(0.9) | 2.7(0.9) | 2.9(0.9) | 3.0(0.9)<sup>*</sup> | 2.9(0.9)<sup>*</sup> | 3.1(0.8)<sup>*</sup> |

BMI=Body mass index; SD=Standard deviation; HCW=Healthcare worker; IMD=Index of Multiple Deprivation; IMD[1-3] indicates high deprivation, IMD [4-7] intermediate, IMD[8-10] low deprivation; age is in years; comorbidity status=at least one comorbidity; for age and BMI the mean and standard deviation are provided, and for categorical variables the absolute number and percent of column total (%).
<sup>1</sup>For the analysis on the healthy lifestyle factor only part of the study population (n=1247) answered the diet questionnaire; the other users were not included in the analysis
\*/= Indicates statistically significant/no statistically significant difference when compared to the case population (Fisher's $p < 0.05$ )

We found significant inverse association between age and post-vaccination infection, which was particularly in older adults (OR=0.96, 95%CI=[0.95-0.97] per year increase in age, p-value<0.0001, see Supplementary Table 2 for detailed results). We also observed a modest but significant positive relationship between BMI and post-vaccination infection especially in younger adults (OR=1.02, CI[1.01-1.03] per unit increase in BMI, p-value<0.0001).

In older adults, frailty (OR=2.78, CI[1.98-3.89], p-value<0.0001) and kidney disease (OR=2.10, CI[1.05-4.21], p-value=0.037) were associated with post-vaccination infection (see Figure 1a and Supplementary Table 3 for detailed results). Post-vaccination infection in frail older adults was not entirely benign, with 30 out of 120 (25%) in this group presenting to hospital. Neither frailty nor any comorbidities were associated with post-vaccination infection in younger adults using p-value<0.05. Sensitivity analysis using IPW for factors influencing vaccination showed consistent results in older adults. In younger adults, decreased odds of post-vaccination were observed in younger adults for frailty, lung disease and diabetes (Supplementary Table 4).

**Figure 1.**
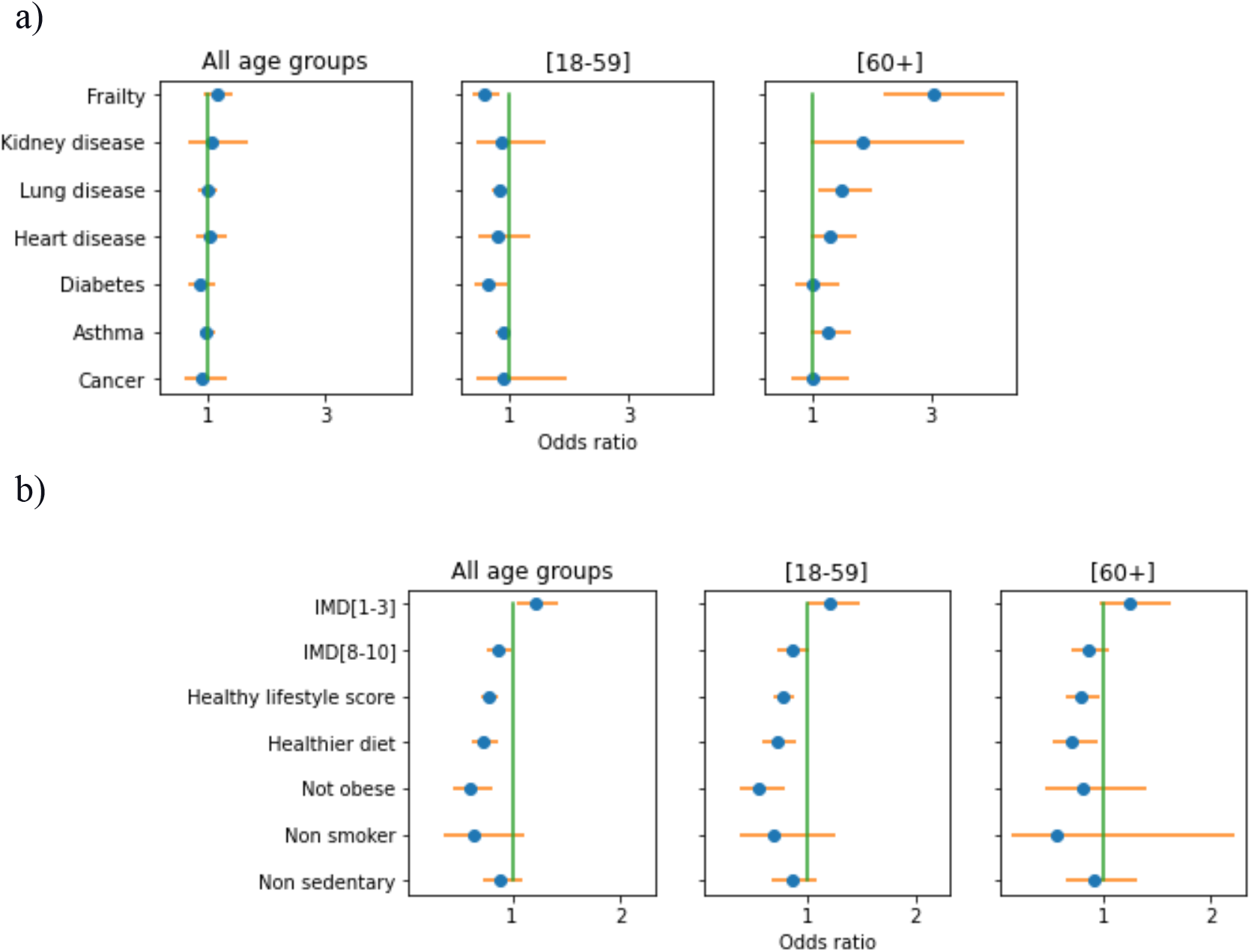
Odds ratio of COVID-19 infection after vaccination: a) Univariate models for frailty and each individual comorbidity, adjusted for age, body mass index (BMI) and sex and stratified by age-group; b) Univariate models for environmental (IMD category), obesity status and healthy lifestyle factors, adjusted for age, BMI, and sex and stratified by age-group. BMI = Body Mass Index; IMD = Index of Multiple Deprivation. IMD[1-3]=high deprivation, IMD[8-10]=low deprivation; reference category for IMD is IMD [4-7] = intermediate category.

Users living in areas of lowest (IMD[8-10]) and highest deprivation (IMD[1-3]) showed, respectively, lower and higher risk compared to the intermediate category (IMD[4-7]) reflecting a progressive increase in risk of post-vaccination infection for individuals living in more deprived areas (IMD[8-10] OR=0.87, CI[0.76-0.98], p-value=0.026; IMD [1-3] OR=1.22, CI[1.04-1.43], p-value=0.013) (Figure 1b and Supplementary Table 5, and 6 for IPW sensitivity checks).

Healthier lifestyle factors were generally associated with lower odds of infection after vaccination regardless of age group. Younger adults with healthier lifestyles had lower odds of post-vaccination infection(OR=0.77, CI[0.68-0.88], per each additional lifestyle factor, p-value<0.0001). The strongest association was seen for body mass index categories, in which individuals with normal weight had lower odds of infection (OR=0.54, CI[0.37-0.78], p-value=0.001). Results were similar in older adults, except obesity, which was no longer significant (Figure 1b and Supplementary Table 5).

In multivariate analysis (Figure 2) diet (all age strata), IMD (younger and all ages) and frailty (older adults) were independently associated with post-vaccination infection (Supplementary Table 7). These findings were consistent in a sensitivity analysis using inverse probability weighting for factors influencing vaccination (Supplementary Tables 8).

**Figure 2.**
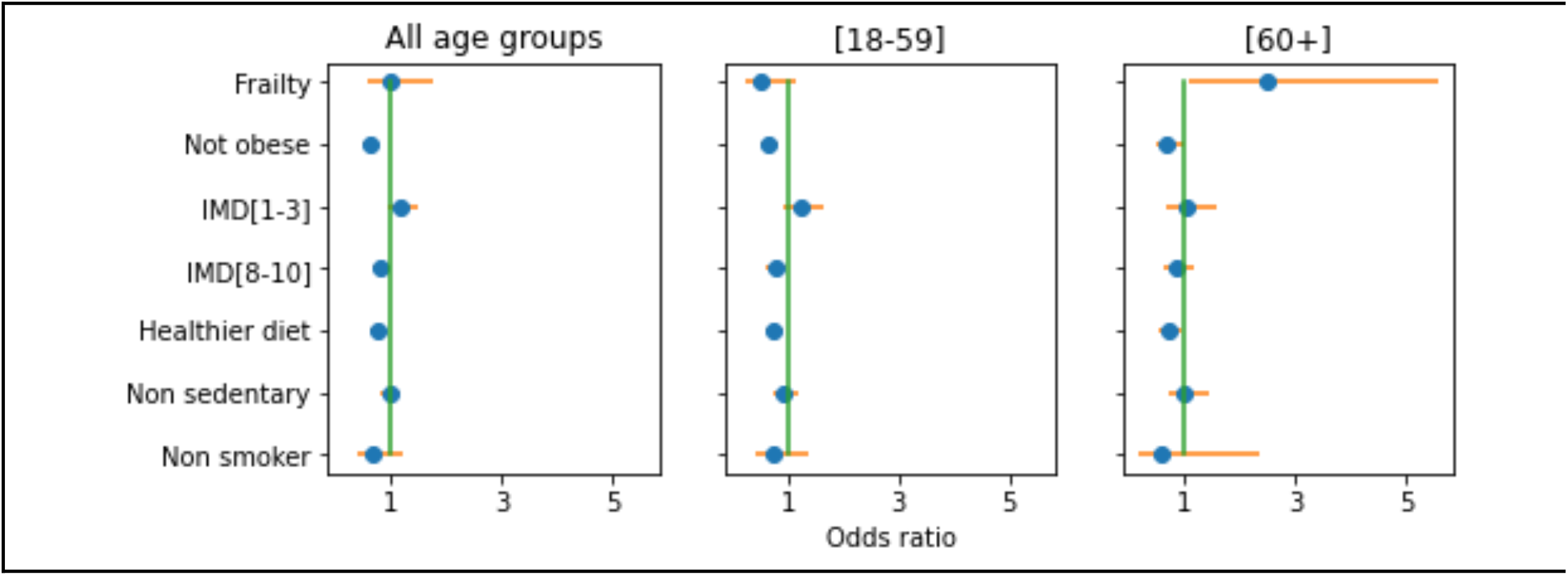
Multivariable analysis of frailty, IMD category, obesity status and healthy lifestyle factors, adjusted for age, BMI and sex. BMI = Body Mass Index; IMD = Index of Multiple Deprivation. IMD[1-3]=high deprivation, IMD[8-10]=low deprivation; reference category for IMD is IMD [4-7] = intermediate category.

### Illness profile in vaccinated and unvaccinated individuals with SARS-CoV-2 infection

A total of 2,188 post-vaccine infected cases were followed-up for at least 14 days post-vaccination (median duration of follow-up: 77 days, IQR 41-101). Matching using nearest Euclidean distance on a number of variables (as per methods) still resulted in a ∼20% preponderance of health care workers in the vaccinated-case group (Table 2). Vaccinated individuals were less likely to have multiple (>5) symptoms in the first week of illness (OR=0.51, CI=[0.42-0.61], p-value<0.0001), present to hospital (OR=0.36, CI[0.28-0.46] p-value<0.0001), and were more likely to be completely asymptomatic (OR=1.72, CI[1.41-2.11], p-value<0.0001 Figure 3, Supplementary Table 9). For older adults, there were lower odds of long-duration symptoms (OR ≥28 days=0.72, CI[0.51-1.00], p-value=0.05).

**Table 2.** Demographics of vaccinated and unvaccinated adults with SARS-CoV 2 infection used for symptom analysis.

|  | Positive group |  |  | Control group of unvaccinated users testing negative (HCW-, sex-, day_of_test-, age-, and BMI-matched) |  |  |
| --- | --- | --- | --- | --- | --- | --- |
|  | All<br>(n=2188, BNT=1363,<br>ChA=748, NS=77) | Younger adults [18-59<br>years]<br>(n=1248, BNT=837,<br>ChA=356, NS=55) | Older adults [60+<br>years]<br>(n=940, BNT=526,<br>ChA=392, NS=22) | All<br>(n=2188) | Younger adults<br>[18-59 years]<br>(n=1260) | Older adults<br>[60+ years]<br>(n=928) |
| **Basic characteristics** |  |  |  |  |  |  |
| Female n(%) | 1533(70.1) | 957(23.3) | 576(38.7) | 1542(70.5)= | 961(23.7)= | 581(37.4)= |
| Age (SD) | 53.0(15.4) | 42.5(9.2) | 66.9(10.0) | 52.8(15.2)* | 42.6(9.2)= | 66.8(9.8)= |
| BMI (SD) | 27.7(7.1) | 27.9(7.4) | 27.3(6.8) | 27.6(7.0)= | 27.9(7.3)= | 27.3(6.5)= |
| HCW n(%) | 624(28.5) | 445(35.7) | 179(19.0) | 489(22.3)* | 351(27.9)* | 138(14.9)* |
| Frailty n (%) | 156(7.1) | 19(1.5) | 137(14.6) | 114(5.2)* | 20(1.6)= | 94(10.1)* |
| Comorbidity status n(%) | 547(25.0) | 255(20.4) | 292(31.1) | 513(23.4)= | 232(18.4)= | 281(30.3)= |
| Asymptomatic infection<br>n(%) | 298(14.2) | 119(10.0) | 179(19.7) | 173(8.8)* | 75(6.6)* | 98(11.8)* |
| Hospitalised n(%) | 104(4.8) | 39(3.2) | 65(7.0) | 239(11.4)* | 76(6.3)* | 163(18.3)* |
| >5 reported symptoms<br>n(%) | 242(15.6) | 139(17.6) | 103(13.6) | 340(29.5)* | 195(31.5)* | 145(27.1)* |
| symptoms<br>lasting ≥28 days<br>n(%) | 134(8.7) | 58(7.3) | 76(10.0) | 124(10.7)* | 42(6.8)= | 82(15.3)* |

BMI=Body mass index; SD=Standard deviation; HCW=Healthcare worker; n=number of individuals; age is in years; Comorbidity status=users with at least one comorbidity; BNT=BNT162b2; ChA=ChAdOx1; NS=not sure; for age and BMI the mean and standard deviation are provided and for categorical variables the absolute value and percentages (%); severity is equal to one if the sum of symptoms experienced in the first week >5.
\*/= Indicates statistically significant/no statistically significant difference when compared to the case population (Fisher's $p < 0.05$ )

**Figure 3.**
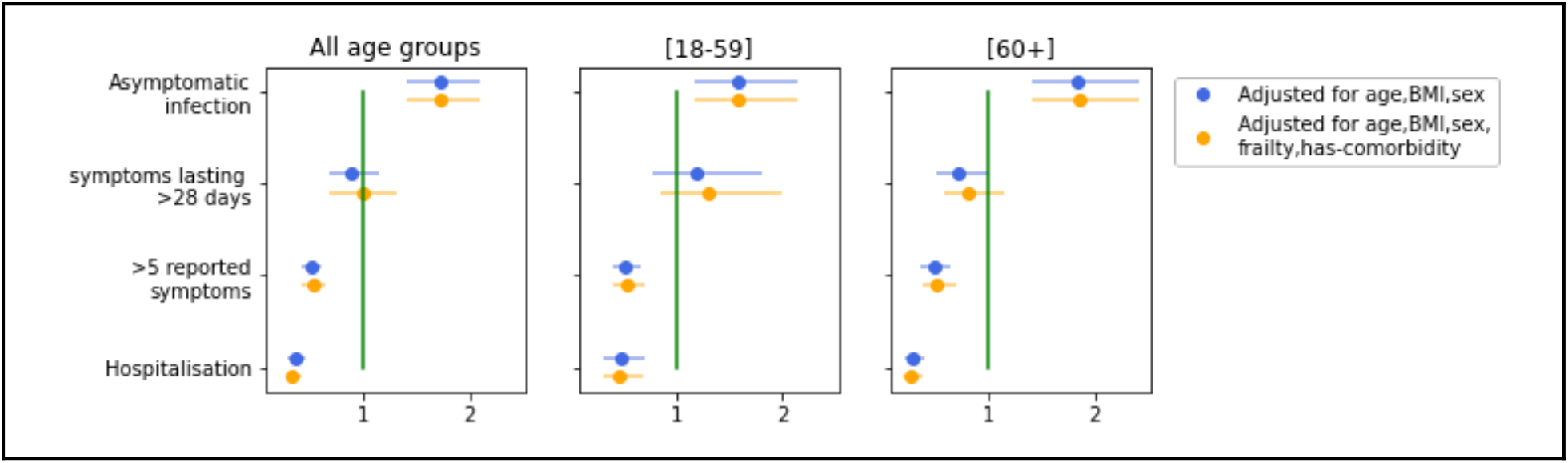
Odds ratio of asymptomatic infection, duration of symptoms > 28 days, severe disease (> 5 reported symptoms during acute infection), and hospitalisation in app participants following vaccination, adjusted by (i) age, BMI, and sex (blue) and (ii) age, BMI, sex, frailty, and comorbidity status (orange).

Analysis of individual symptoms (Figure 4 and Supplementary Table10), showed that vaccination was associated with lower symptom reporting for almost all symptoms across all age groups, with the exceptions of sneezing (sternutation), which was more common in vaccinated individuals (OR=1.24 95%CI [1.05-1.46], p-value=0.01,) and shortness of breath, earache and swollen glands that were no different between study groups. Results remained similar after adjustment for comorbidities and frailty (Supplementary Tables 11 and 12).

**Figure 4.**
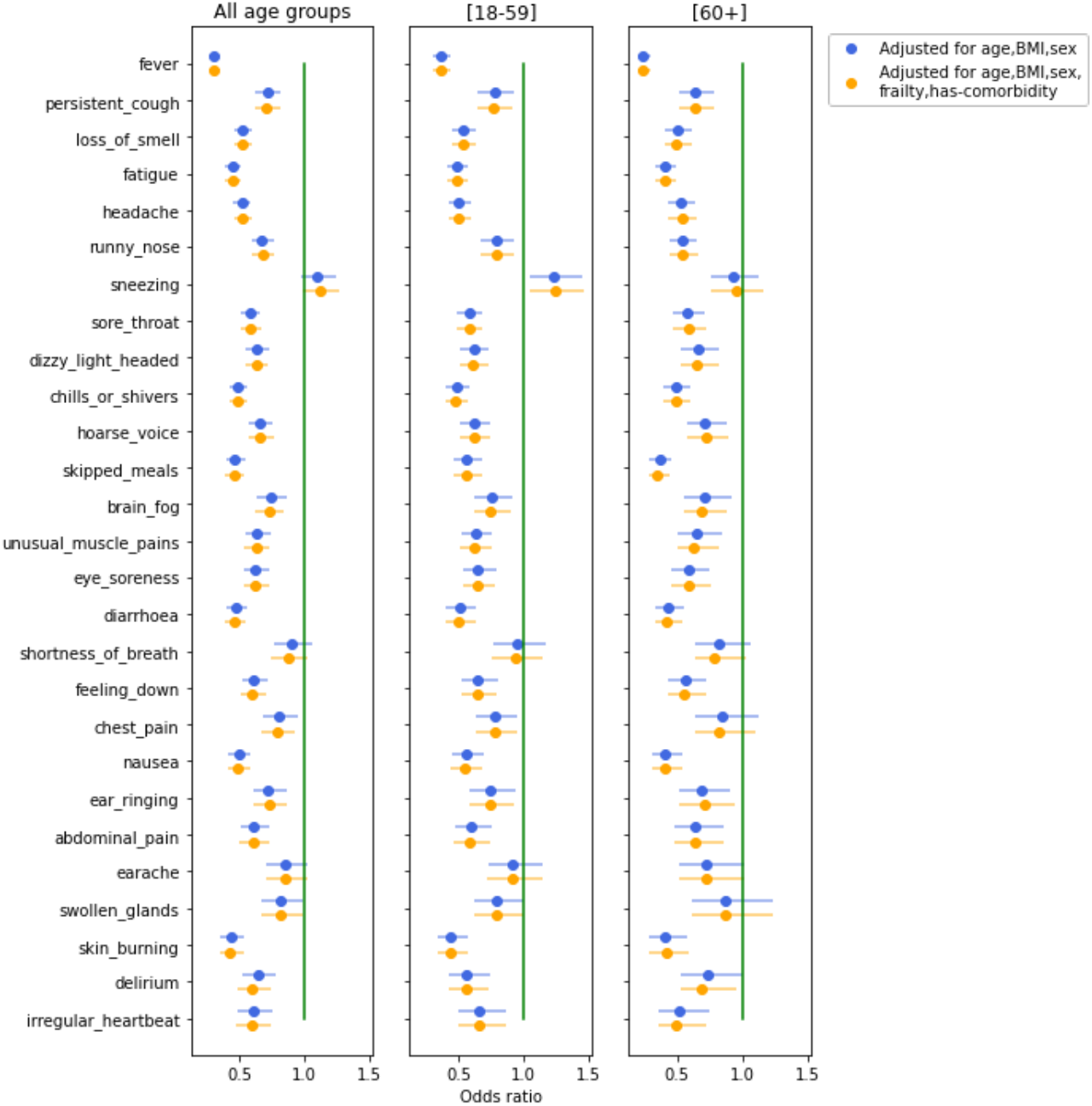
Odds Ratio of individual symptoms in vaccinated versus unvaccinated app participants with reported SARS-CoV-2 infection, adjusted by (i) age, BMI, and sex (blue) and (ii) age, BMI, sex, frailty, and comorbidity status (orange).

## Discussion

As more countries vaccinate their populations against COVID-19, there is growing interest in understanding risk factors for and characteristics of post-vaccination infection, to guide health policies and resource planning. Here, we present data on 2,394 community-based adults in the UK with test-confirmed SARS-CoV-2 infection more than 14 days after first vaccination with BNT162b2 or ChAdOx1, when immunity is starting to develop (31) and infection is unlikely to be due to exposure peri-vaccination (e.g. during travel to the vaccination centre).

Frail individuals had higher odds of post-vaccine SARS-CoV-2 infection than matched controls, highlighting need for ongoing caution in this vulnerable group. The association was consistent in sensitivity analysis using inverse probability weighting for factors influencing vaccination, and when adjusting for potential confounders including local area deprivation and lifestyle. This may reflect increased exposure: unlike robust older adults, frailer adults may require carer visits or to attend healthcare facilities. Frail adults in long-term care facilities are at particular risk of transmission of respiratory illness, and have been disproportionately affected throughout the pandemic (32). Another explanation may be altered immune function (“immunosenescence”), a well-established feature of physiological ageing (33). Furthermore, immunosenescence may explain previously observed ageing-associated decline in immunogenicity following other vaccinations (34). The increased odds of post-vaccine infection in frailer adults may be compounded by more severe outcomes of COVID-19 infection in this group, including delirium (24) and death (17); indeed this study a quarter of frail older adults testing positive after vaccination required assessment in hospital. NICE recommends systematic frailty assessment in acute settings, (https://www.nice.org.uk/guidance/NG159); this could extend to community settings to facilitate differential, targeted re-vaccination scheduling, appropriate isolation precautions, case detection, testing, and proactive care. Research on augmenting immunogenicity in this group is urgently needed; for example, on impact and timing of booster vaccinations.

We found an inverse association of age on odds of post-vaccination infection, especially in older adults. This is consistent with previous studies in non-vaccinated individuals showing lower antibody seroprevalence in older adults (35), likely reflecting shielding in this age-group in accordance with classification of over 70s as clinically vulnerable (36). We found evidence that kidney disease may increase odds of infection post-vaccination. This is potentially important, as patients with kidney disease were under-represented in the Phase 2 and Phase 3 trials of vaccines (37) and may reflect increased exposure (e.g. attending for dialysis etc), or impaired immunogenicity in these individuals as observed for other infections (38, 39). In our cohort, neither cancer nor other comorbidities were significantly associated with an increased odds of infection. While this is reassuring, given that many of these comorbidities confer higher risk of severe disease, hospitalisation, mechanical ventilation, and mortality from COVID-19 (16, 41), as with age, ongoing shielding behaviours may be influencing our results.

Greater area level deprivation was associated with risk of post-vaccination SARS-CoV-2 infection consistent with findings from the pre-vaccination era (42). This association persisted following adjustment for compliance with infection control guidance (mask-wearing). Associated factors including higher density and more ethnically diverse populations are also associated with higher mortality from COVID-19 (43, 44). Individuals in more deprived areas may have lower vaccination coverage for COVID-19 (45), and our finding may reflect increased viral transmission in these areas. Our findings suggest that health policies to mitigate infection will need to be targeted to these areas.

Conversely, individuals reporting healthier lifestyles, in particular healthier diet, had lower risk of infection post-vaccination as did those without obesity, which was associated with adverse outcomes pre-vaccination. This suggests that immune responses post-vaccination may be influenced by diet quality and obesity, although unadjusted confounding remains a possibility.

Almost all individual symptoms of COVID-19 disease were less common in the vaccinated vs. unvaccinated populations with SARS-CoV-2 infection, and more people were completely asymptomatic in the vaccinated group. The exception was sternutation (sneezing), reported more commonly in younger adults who contracted infection post-vaccination. We could not find previous reports of this for other respiratory illnesses, but sneezing is a well-recognised symptom of common upper respiratory infections and allergy, prompted by nasal mucosa antigen irritation. Activation of the immune system can heighten the sneeze response to an antigen through inflammatory cascades including cytokine production and neuropeptides (46). Immune system ‘priming’ with vaccination might lead to an increase in sneezing in response to the SARS-CoV-2 antigen; an appropriate adaptive response to clear the virus. However, sneezing generates aerosols; potentially of importance for viral transmission in the post-vaccine era but is not currently a core symptom triggering testing. Put together with the increase in asymptomatic cases, this underlines the need for those interacting with unvaccinated or vulnerable groups (e.g. health and social care workers) to continue to test regularly for SARS-CoV-2.

We found lower severity of COVID-19 disease (both number of symptoms in the first week of infection, and need for hospitalisation) in vaccinated compared to unvaccinated individuals, and in older adults, less risk of Long-COVID. We have previously shown that experiencing more than 5 symptoms in the acute period was associated with severity of disease (19) and duration of symptoms (20). These findings suggest that severe acute disease, hospitalisation rates and Long-COVID prevalence will fall in the post-vaccination era, and those who need hospital are likely to be older people with frailty.

### Strengths and Limitations

This study used data from a large population of individuals reporting on a mobile application. This population, while large, was disproportionately female and under-represented individuals of lower socio-economic status as indicated by the skew toward people living in less deprived areas (Table 1).

Information was self-reported and therefore recording of comorbidities and test results may not be completely accurate. However, previous data from this study have concurred well with population-based COVID-19 studies (47), including the influence of socio-demographic factors (42). A strength of the mobile data collection method is the ability to collect daily information prospectively, on a comprehensive set of symptoms, allowing analysis of both individual symptoms and overall illness duration. We acknowledge also that by virtue of data censoring dates, symptom duration may be underestimated in both cases and controls, as some individuals only had two weeks of logging after their positive test result.

The design of our study, including matching cases and controls for health-care worker status and time of infection, reduces potential for bias, although small differences between the groups remained on matched variables. Risk of reporting a positive SARS-CoV-2 test is higher amongst frontline healthcare workers vs. the general population (14), reflecting exposure; and appropriately, healthcare workers were prioritised for vaccination in the UK (48). Our data suggests risk of post-vaccination SARS-CoV-2 infection is reduced in older age groups. In order to examine the effect of age on post-vaccination infection, we did not match CG1 by age. However, age was included as a covariate in all analyses other than that looking at effects of age itself, and stratified analyses are presented in two age-groups. While vaccination itself might be considered a potential index event bias, the population of interest in this study is the vaccinated population, and should not be construed as applying to those unvaccinated. Nevertheless, we examined and found no evidence of event bias based on probability of being vaccinated.

Frailty was assessed with the PRISMA-7 questionnaire for app usage. This assessment correlates well with other frailty measures (49) and has the advantage of focusing on functional consequences of frailty, not routinely captured in health records. However, PRISMA-7 has only been validated in older adults; results in younger adults should be interpreted cautiously (23).

Finally, this study was conducted at the beginning of the post-vaccination period, at a time when incidence of SARS-CoV-2 infection in the UK was rapidly falling. Concurring with recently published ISARIC data, we saw declining incidence of new infections with time after vaccination (12), which may reflect both increasing immunity and falling incidence in the population. Our findings may not apply at all time points post-vaccination. Lastly, the small number of individuals who had received a second vaccination precluded study of post-vaccination infection after more than one dose.

## Conclusions

We investigated factors associated with increased risk of infection after vaccination and found a substantial increased risk in frail older adults and in individuals living in more deprived areas, and a lower risk of infection in non-obese people and those who reported better diet quality. We found most symptoms post-vaccination were reported less in vaccinated people, except sneezing. Need for hospital assessment was less, burden of acute symptoms was lower and for older adults, risk of prolonged illness was lower. Our findings may inform policy in the post-vaccination era, in particular to protect frail older adults, and those individuals living in areas of higher relative deprivation. This research suggests focused infection control measures should continue to be in place for these populations, to minimise their risk of COVID-19, while strategies, such as booster vaccination, are explored.

## Data Availability

Data used in this study are available to bona fide researchers through UK Health
Data Research at https://web.www.healthdatagateway.org/dataset/fddcb382-3051-4394-8436-b92295f14259/

## Acknowledgements

Zoe Global provided in-kind support for all aspects of building, running and supporting the app and service to all users worldwide. This work is supported by the UK Department of Health via the National Institute for Health Research (NIHR) comprehensive Biomedical Research Centre award to Guy’s & St Thomas’ NHS Foundation Trust in partnership with King’s College London and King’s College Hospital NHS Foundation Trust, and a grant to ZOE Globals, as well as by the Wellcome EPSRC Centre for Medical Engineering at King’s College London (WT 203148/Z/16/Z). This work was further supported by the UK Research and Innovation London Medical Imaging & Artificial Intelligence Centre for Value-Based Healthcare. Investigators also received support from the Wellcome Trust, Medical Research Council (MRC), British Heart Foundation (BHF), Alzheimer’s Society, European Union, NIHR, Chronic Disease Research Foundation (CDRF) and the NIHR-funded BioResource, Clinical Research Facility and Biomedical Research Centre (BRC) based at GSTT NHS Foundation Trust in partnership with KCL. S.O. was supported by the French government, through the 3IA Côte d’Azur Investments in the Future project managed by the National Research Agency (ANR) with the reference number ANR-19-P3IA-0002. A.T.C. was supported by a Stuart and Suzanne Steele MGH Research Scholar Award and by the Massachusetts Consortium on Pathogen Readiness (MassCPR) and M. Schwartz and L. Schwartz. J.M. was partially supported by the European Commission Horizon 2020 program (H2020-MSCA-IF-2015-703787).

---

**Supplementary Table 1.** List of self-reported symptoms and corresponding questions used in the reporting app.

| Symptoms | Questions |
| --- | --- |
| fever | Do you have a fever or feel too hot? |
| chills or shivers | Do you feel chills or shivers (feel too cold)? |
| persistent cough | Persistent cough (coughing a lot for more than an hour or 3 or more coughing episodes in 24 hours) |
| fatigue | Are you experiencing unusual fatigue? |
| shortness of breath | Shortness of breath or trouble breathing |
| loss of smell | Loss of smell / taste |
| hoarse voice | Unusually hoarse voice |
| chest pain | Unusual chest pain or tightness in your chest |
| abdominal pain | Unusual abdominal pain or stomach ache |
| diarrhoea | Diarrhoea |
| delirium | Confusion, disorientation or drowsiness |
| eye soreness | Do your eyes have any unusual eye-soreness or discomfort (e.g. light sensitivity, excessive tears, or pink/red eye)? |
| skipped meals | Skipping meals |
| headache | Headache |
| nausea | Nausea or vomiting |
| dizzy light headed | Dizziness or light-headedness |
| sore throat | Sore or painful throat |
| unusual muscle pains | Unusual strong muscle pains or aches |
| red welts on face or lips | Raised, red, itchy welts on the skin or sudden swelling of the face or lips |
| blisters on feet | Red/purple sores or blisters on your feet, including your toes |
| typical hayfever | Increase in your usual allergy symptoms |
| rash | Rash on your arms or torso |
| skin burning | Strange, unpleasant sensations in your skin like pins & needles or burning |
| hair loss | Unusual hair loss |
| feeling down | Feeling down, depressed or hopeless |
| brain fog | Loss of concentration or memory (brain fog) |
| runny nose | Runny nose |
| sneezing | Sneezing more than usual |
| earache | Earache |
| ear ringing | Ringing in your ears |
| swollen glands | Swollen neck glands |
| irregular heartbeat | Unusually fast or irregular heartbeat (palpitations) |

**Supplementary Table 2.**
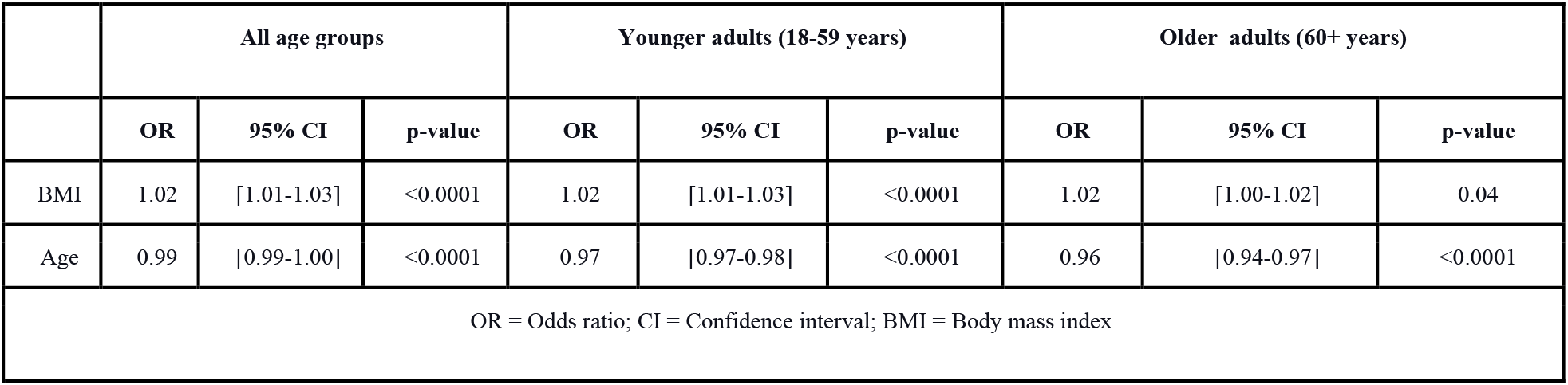
Multivariable analysis of age and body mass index (BMI), adjusted by sex.

**Supplementary Table 3.**
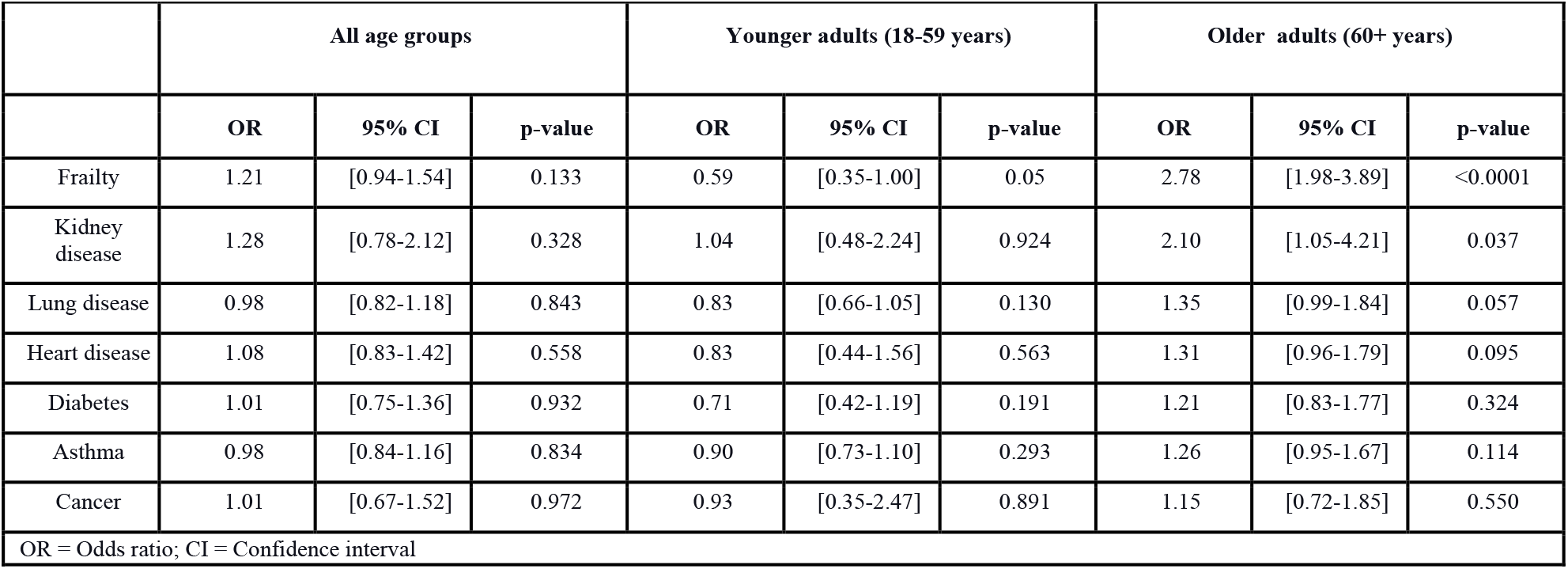
Univariate analysis of frailty status and each comorbidity, adjusted by age, BMI, and sex.

|  | All age groups |  |  | Younger adults (18-59 years) |  |  | Older adults (60+ years) |  |  |
| --- | --- | --- | --- | --- | --- | --- | --- | --- | --- |
|  | OR | 95% CI | p-value | OR | 95% CI | p-value | OR | 95% CI | p-value |
| Frailty | 1.21 | [0.94-1.54] | 0.133 | 0.59 | [0.35-1.00] | 0.05 | 2.78 | [1.98-3.89] | <0.0001 |
| Kidney disease | 1.28 | [0.78-2.12] | 0.328 | 1.04 | [0.48-2.24] | 0.924 | 2.10 | [1.05-4.21] | 0.037 |
| Lung disease | 0.98 | [0.82-1.18] | 0.843 | 0.83 | [0.66-1.05] | 0.130 | 1.35 | [0.99-1.84] | 0.057 |
| Heart disease | 1.08 | [0.83-1.42] | 0.558 | 0.83 | [0.44-1.56] | 0.563 | 1.31 | [0.96-1.79] | 0.095 |
| Diabetes | 1.01 | [0.75-1.36] | 0.932 | 0.71 | [0.42-1.19] | 0.191 | 1.21 | [0.83-1.77] | 0.324 |
| Asthma | 0.98 | [0.84-1.16] | 0.834 | 0.90 | [0.73-1.10] | 0.293 | 1.26 | [0.95-1.67] | 0.114 |
| Cancer | 1.01 | [0.67-1.52] | 0.972 | 0.93 | [0.35-2.47] | 0.891 | 1.15 | [0.72-1.85] | 0.550 |

OR = Odds ratio; CI = Confidence interval

**Supplementary Table 4.** Sensitivity analysis of frailty and each comorbidity using Inverse Probability Weighting (IPW) for probability of vaccination. Each univariate analysis is adjusted by age, BMI, and sex.

|  | All age groups |  |  | Younger adults (18-59 years) |  |  | Older adults (60+ years) |  |  |
| --- | --- | --- | --- | --- | --- | --- | --- | --- | --- |
|  | OR | 95% CI | p-value | OR | 95% CI | p-value | OR | 95% CI | p-value |
| Frailty | 1.15 | [0.93-1.43] | 0.189 | 0.55 | [0.37-0.82] | 0.003 | 3.06 | [2.20-4.25] | <0.0001 |
| Kidney disease | 1.06 | [0.67-1.67] | 0.818 | 0.84 | [0.44-1.61] | 0.607 | 1.84 | [0.95-3.55] | 0.07 |
| Lung disease | 0.99 | [0.86-1.15] | 0.933 | 0.81 | [0.68-0.97] | 0.018 | 1.5 | [1.11-2.02] | 0.008 |
| Heart disease | 1.05 | [0.82-1.34] | 0.718 | 0.8 | [0.48-1.34] | 0.396 | 1.3 | [0.96-1.75] | 0.084 |
| Diabetes | 0.87 | [0.67-1.14] | 0.327 | 0.63 | [0.41-0.96] | 0.031 | 1.01 | [0.71-1.45] | 0.939 |
| Asthma | 0.98 | [0.86-1.12] | 0.748 | 0.89 | [0.76-1.03] | 0.116 | 1.27 | [0.97-1.66] | 0.084 |
| Cancer | 0.89 | [0.60-1.32] | 0.56 | 0.9 | [0.42-1.95] | 0.794 | 1.01 | [0.64-1.62] | 0.951 |

OR = Odds ratio; CI = Confidence interval

**Supplementary Table 5.**
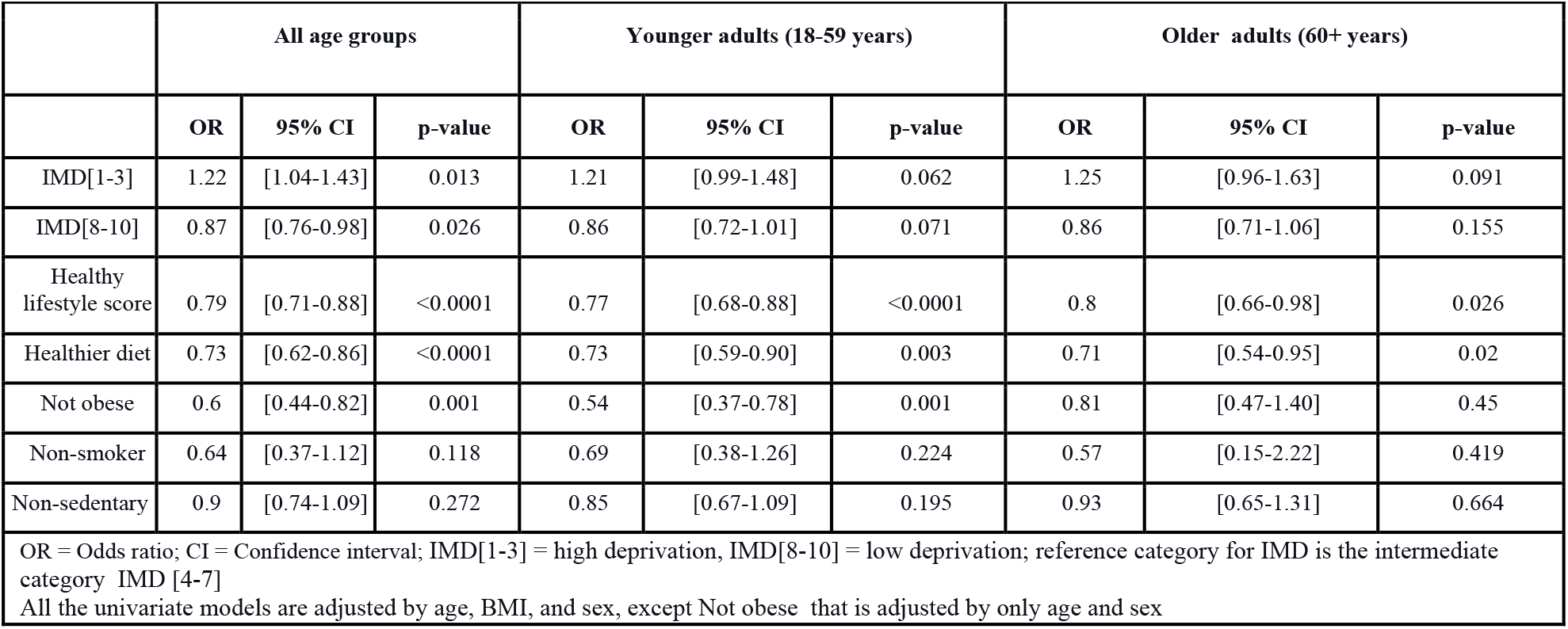
Univariate analysis of Index of Multiple Deprivation (IMD) category, obesity status and healthy lifestyle factors.

**Supplementary Table 6.**
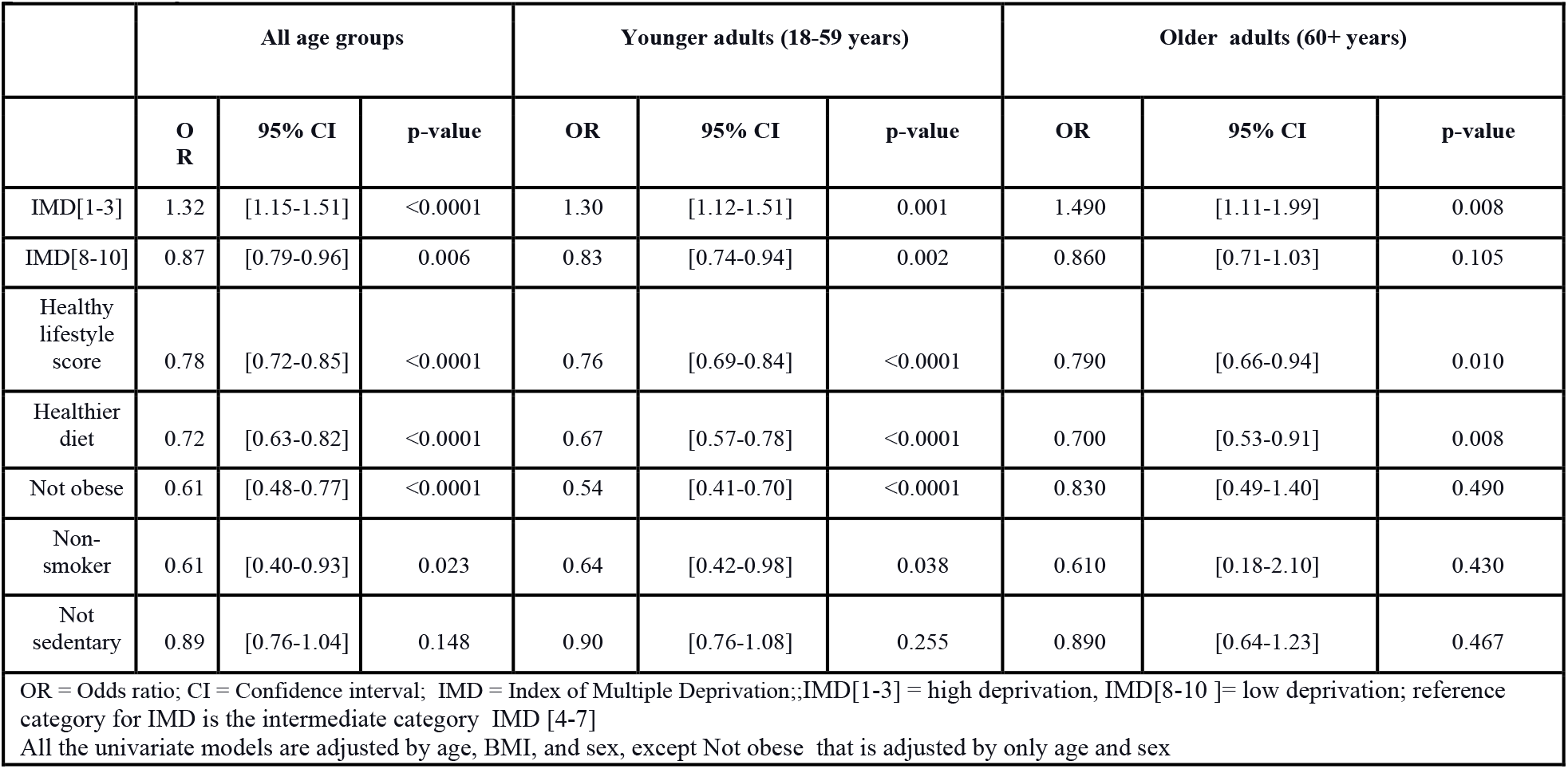
Sensitivity analysis of univariate analyses for IMD categories, obesity status, and healthy lifestyle factors, using Inverse Probability Weighting (IPW) for probability of vaccination.

**Supplementary Table 7.**
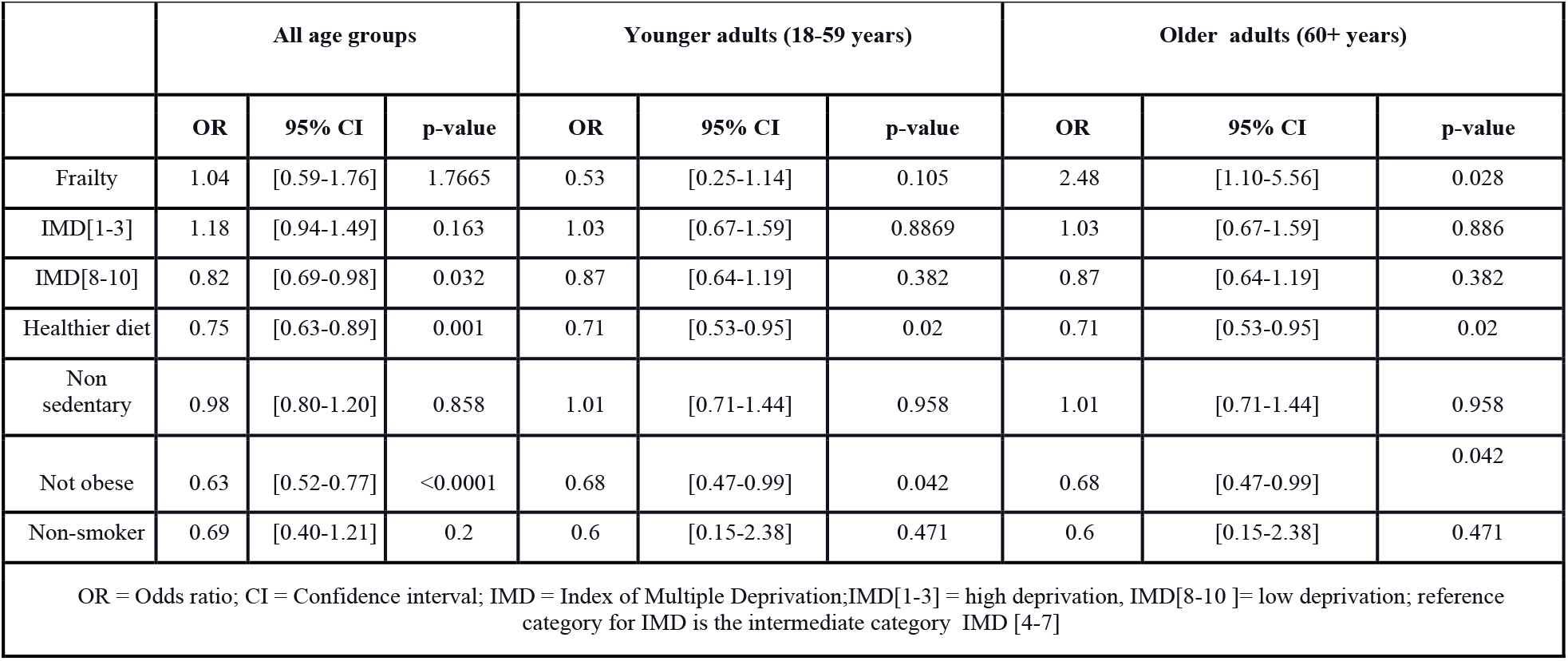
Multivariable analysis of frailty, IMD category, obesity status and healthy lifestyle factors, adjusted for age and sex.

**Supplementary Table 8.**
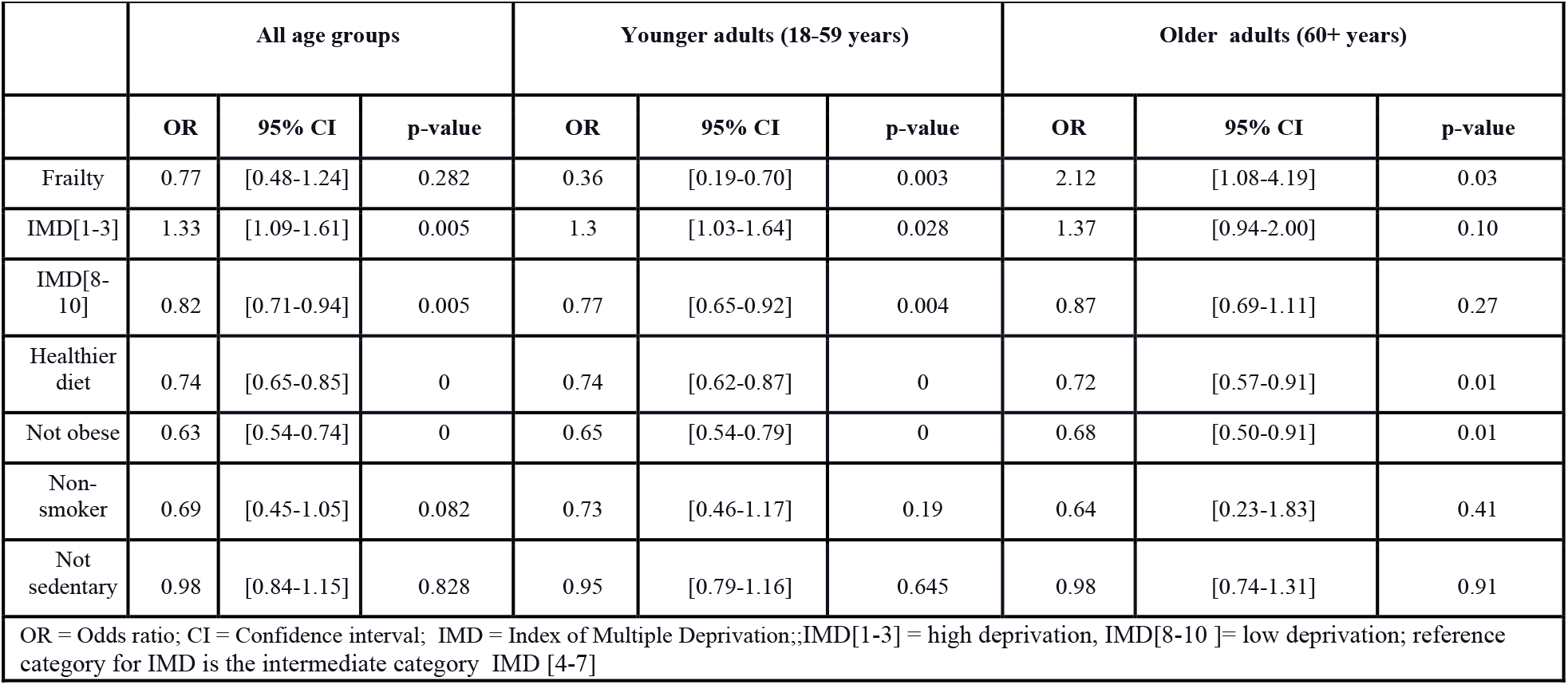
Sensitivity analysis of the multivariable model for frailty, IMD categories, obesity status, and healthy lifestyle factors, using Inverse Probability Weighting (IPW) for probability of vaccination. The analysis is adjusted by age and sex.

**Supplementary Table 9.**
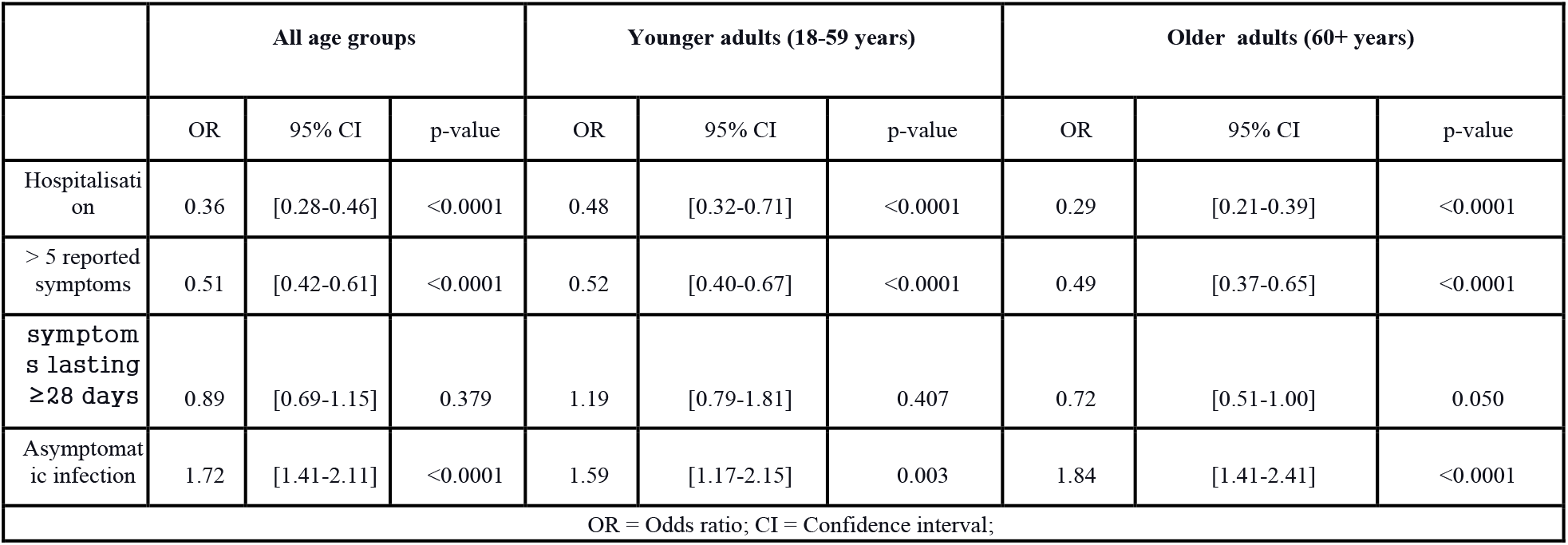
Univariate analysis assessing the probability of asymptomatic infection, severe disease (> 5 reported symptoms during acute infection), hospitalisation and duration of symptoms > 28 days in app participants following vaccination, adjusted by age, BMI, and sex.

|  | All age groups |  |  | Younger adults (18-59 years) |  |  | Older adults (60+ years) |  |  |
| --- | --- | --- | --- | --- | --- | --- | --- | --- | --- |
|  | OR | 95% CI | p-value | OR | 95% CI | p-value | OR | 95% CI | p-value |
| Hospitalisation | 0.36 | [0.28-0.46] | <0.0001 | 0.48 | [0.32-0.71] | <0.0001 | 0.29 | [0.21-0.39] | <0.0001 |
| > 5 reported symptoms | 0.51 | [0.42-0.61] | <0.0001 | 0.52 | [0.40-0.67] | <0.0001 | 0.49 | [0.37-0.65] | <0.0001 |
| symptoms lasting ≥28 days | 0.89 | [0.69-1.15] | 0.379 | 1.19 | [0.79-1.81] | 0.407 | 0.72 | [0.51-1.00] | 0.050 |
| Asymptomatic infection | 1.72 | [1.41-2.11] | <0.0001 | 1.59 | [1.17-2.15] | 0.003 | 1.84 | [1.41-2.41] | <0.0001 |
| OR = Odds ratio; CI = Confidence interval; |  |  |  |  |  |  |  |  |  |

**Supplementary Table 10.** Univariate analysis assessing the probability of experiencing each symptom in app participants following vaccination, adjusted by age, BMI, sex.

|  | All age groups |  |  | Younger adults (18-59 years) |  |  | Older adults (60+ years) |  |  |
| --- | --- | --- | --- | --- | --- | --- | --- | --- | --- |
|  | OR | 95% CI | p-value | OR | 95% CI | p-value | OR | 95% CI | p-value |
| Fever | 0.31 | [0.27-0.36] | <0.0001 | 0.37 | [0.30-0.44] | <0.0001 | 0.24 | [0.19-0.30] | <0.0001 |
| Persistent cough | 0.72 | [0.63-0.82] | <0.0001 | 0.78 | [0.65-0.92] | 0.004 | 0.64 | [0.52-0.78] | <0.0001 |
| Loss of smell | 0.53 | [0.46-0.60] | <0.0001 | 0.54 | [0.46-0.64] | <0.0001 | 0.5 | [0.40-0.61] | <0.0001 |
| Fatigue | 0.45 | [0.40-0.51] | <0.0001 | 0.49 | [0.41-0.58] | <0.0001 | 0.4 | [0.33-0.49] | <0.0001 |
| Headache | 0.52 | [0.46-0.59] | <0.0001 | 0.5 | [0.42-0.60] | <0.0001 | 0.53 | [0.43-0.64] | <0.0001 |
| Runny nose | 0.68 | [0.60-0.76] | <0.0001 | 0.79 | [0.67-0.93] | 0.005 | 0.54 | [0.44-0.65] | <0.0001 |
| Sneezing | 1.1 | [0.97-1.25] | 0.136 | 1.24 | [1.05-1.46] | 0.011 | 0.92 | [0.76-1.13] | 0.449 |
| Sore throat | 0.58 | [0.51-0.66] | <0.0001 | 0.58 | [0.49-0.69] | <0.0001 | 0.58 | [0.47-0.71] | <0.0001 |
| Dizziness or lightheadedness | 0.63 | [0.55-0.73] | <0.0001 | 0.62 | [0.52-0.74] | <0.0001 | 0.66 | [0.53-0.82] | <0.0001 |
| Chills or shivers | 0.49 | [0.42-0.56] | <0.0001 | 0.49 | [0.40-0.59] | <0.0001 | 0.49 | [0.39-0.60] | <0.0001 |
| Hoarse voice | 0.66 | [0.57-0.76] | <0.0001 | 0.62 | [0.52-0.75] | <0.0001 | 0.71 | [0.57-0.89] | 0.003 |
| Skipped meals | 0.47 | [0.40-0.55] | <0.0001 | 0.57 | [0.46-0.69] | <0.0001 | 0.36 | [0.29-0.46] | <0.0001 |
| Brain fog | 0.74 | [0.64-0.86] | <0.0001 | 0.76 | [0.63-0.91] | 0.003 | 0.72 | [0.56-0.92] | 0.009 |
| Unusual muscle pains | 0.64 | [0.55-0.74] | <0.0001 | 0.63 | [0.52-0.76] | <0.0001 | 0.65 | [0.50-0.84] | 0.001 |
| Eye soreness | 0.63 | [0.54-0.73] | <0.0001 | 0.65 | [0.54-0.79] | <0.0001 | 0.58 | [0.45-0.75] | <0.0001 |

**Supplementary Table 11.**
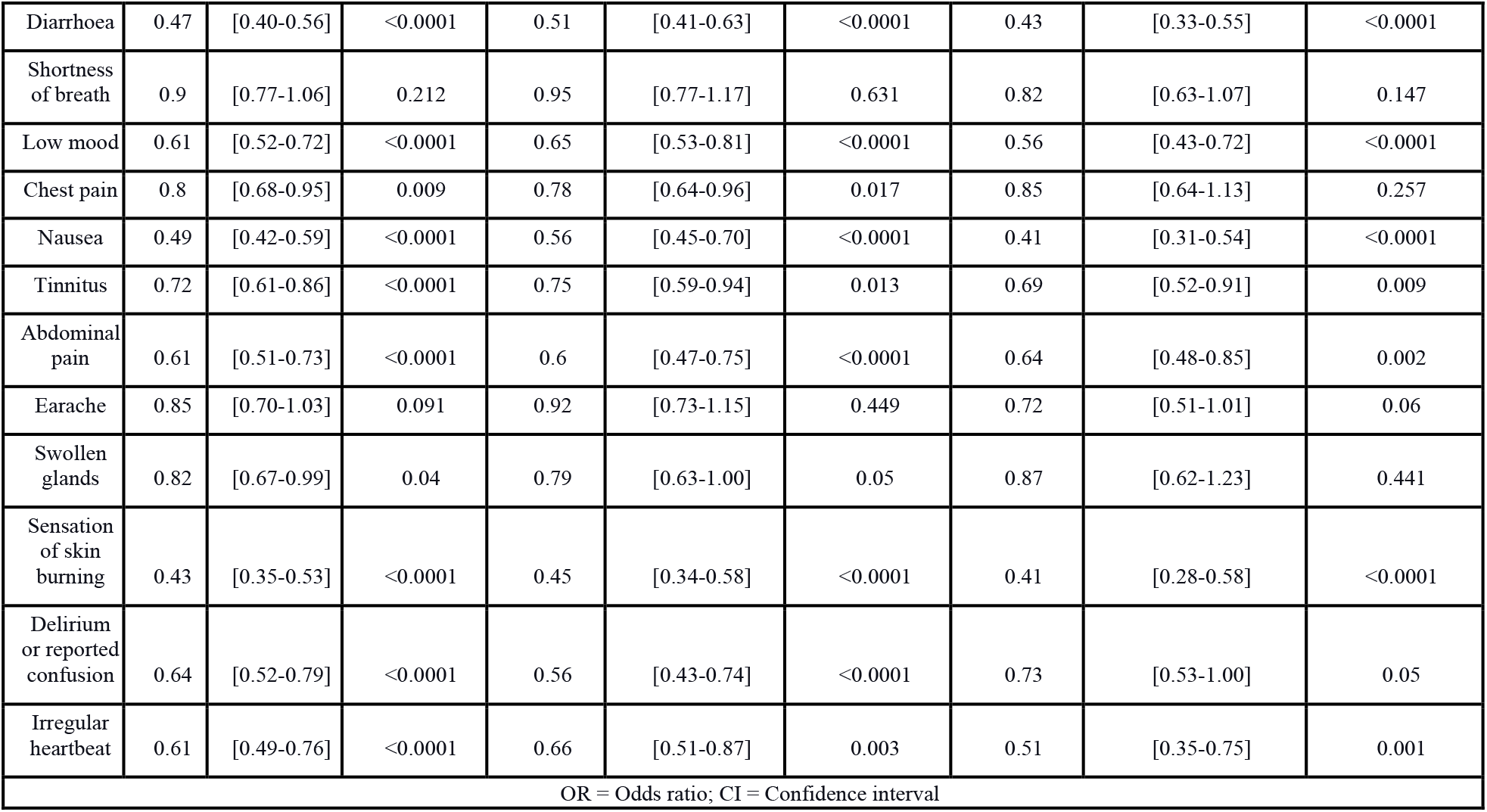
Univariate analysis assessing the probability of asymptomatic infection, severe disease (> 5 reported symptoms during acute infection), hospitalisation, and duration of symptoms > 28 days in app participants following vaccination, adjusted by age, BMI, sex, frailty, and comorbidity status.

**Supplementary Table 12.**
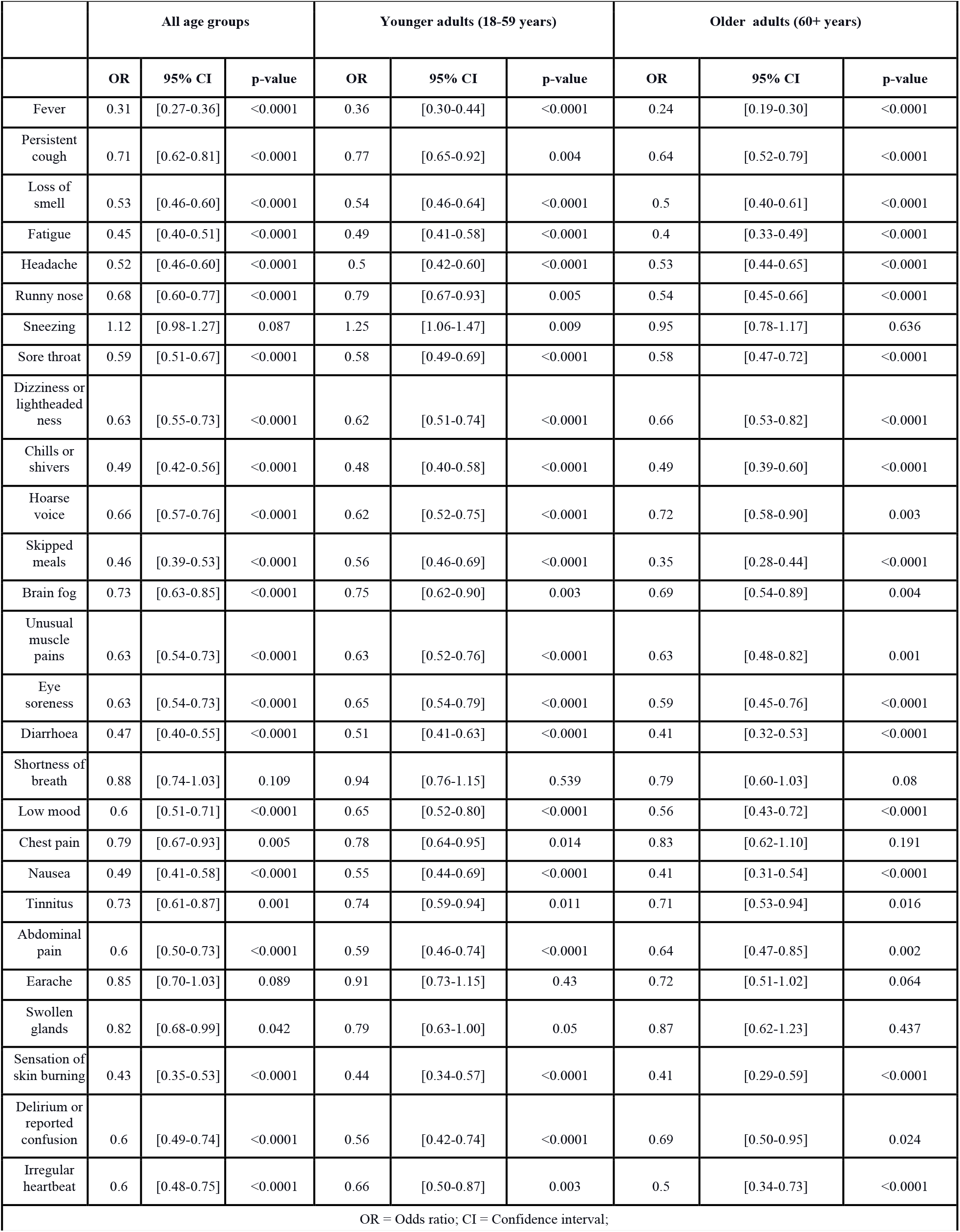
Univariate analysis assessing the probability of experiencing each symptom in app participants following vaccination, adjusted by age, BMI, sex, frailty, and comorbidity status.

**Supplementary Figure 1.**
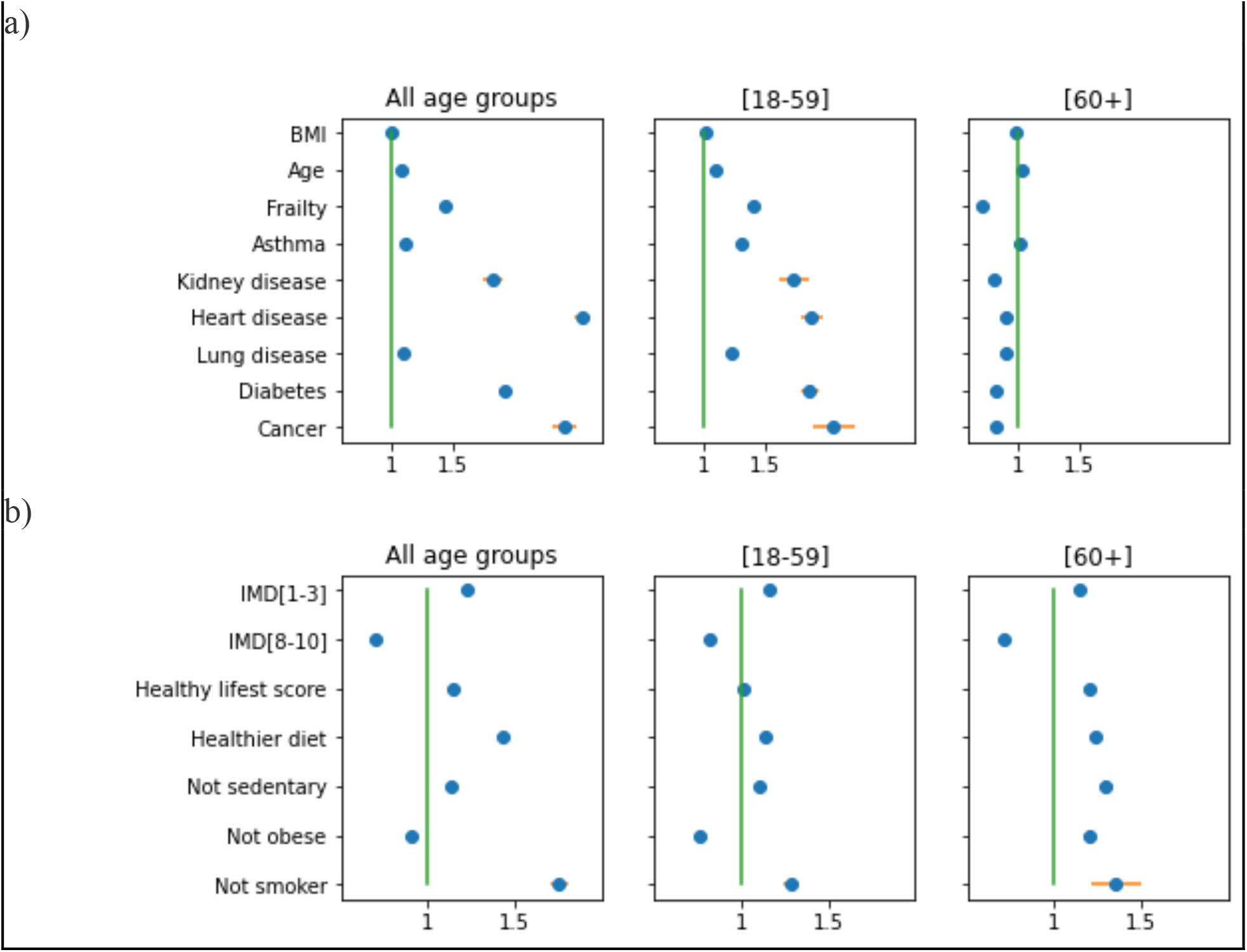
Odds ratio of receiving the vaccine for all users who have recorded at least one COVID-19 test since 8th of December 2020. Univariate analysis for a) age, body mass index (BMI), comorbidity status, and frailty; and b) Index of Multiple Deprivation category and healthy lifestyle factors, adjusted for age, BMI, and sex and stratified by age group. IMD = Index of Multiple Deprivation;;IMD[1-3] = high deprivation, IMD[8-10]= low deprivation; reference category for IMD is the intermediate category IMD [4-7]

**Supplementary Figure 2.**
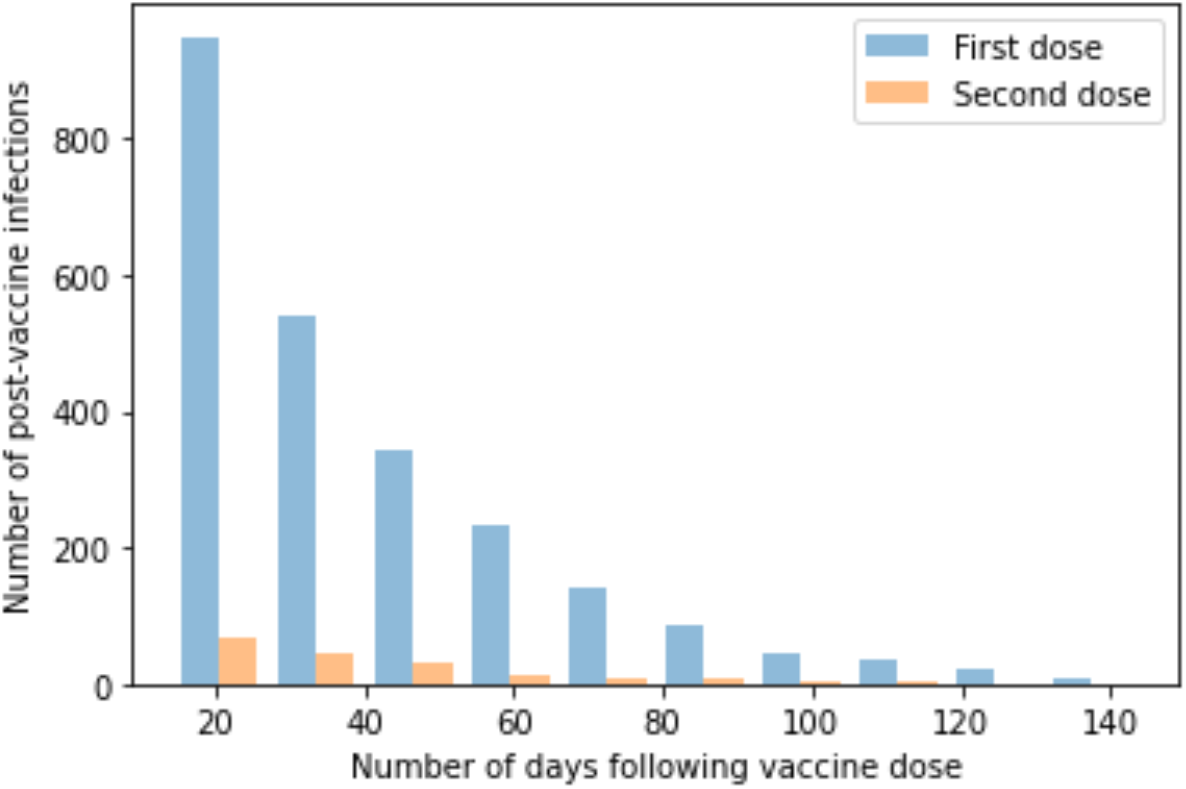
Histogram illustrating number of reported positive tests against number of days from first (blue bar) and second (orange bar) vaccine dose and for COVID-19 infection. NB these data are not adjusted for incidence of infection which changed over the same time period in the UK (29).

**Supplementary Figure 3.**
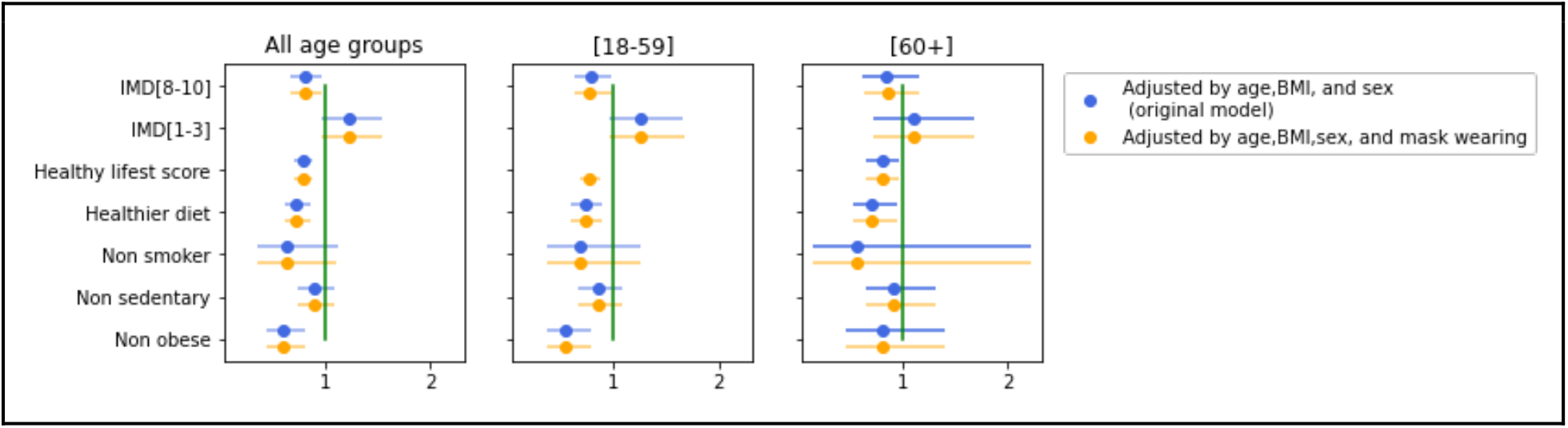
Comparison between the original univariate model, adjusted for age, body mass index, and sex and the model adjusted for i) age, BMI, sex (blue);and ii) age, BMI, sex, and mask-wearing (orange).

## Supplementary Methods

### Dietary Assessment

Diet was assessed using information obtained from an amended version of the Leeds Short Form Food Frequency Questionnaire that included 27 food items (50). Participants were asked how often on average they had consumed one portion of each food in a typical week during the month just prior (July 2020) to when they filled out the diet and lifestyle questionnaire. The responses had eight frequency categories ranging from “rarely or never” to “five or more times per day”. A healthy diet pattern was ascertained using the Diet Quality Score (DQS), a validated score for adherence to UK dietary guidelines (50). The DQS was computed from five broad categories including fruits, vegetables, total fat, oily fish, and non-milk extrinsic sugars. Each component was scored from 1 (unhealthiest) to 3 (healthiest) points, with intermediate values scored proportionally. All component scores were summed to obtain a total score ranging from 5 (lowest diet quality) to 15 (highest) points. We defined a healthier diet pattern as a DQS in the top quartile of the score distribution (score>=12 points). To generate the lifestyle scores, the participants received 1 point for each healthy lifestyle factor. The sum of these four scores together gave a healthy lifestyle score ranging from 0 to 4, with higher scores indicating a healthier lifestyle.

